# Seroprevalence of SARS-CoV-2 during pregnancy and associated outcomes: results from an ongoing prospective cohort study, New York City

**DOI:** 10.1101/2021.02.01.21250943

**Authors:** Nina M. Molenaar, Anna-Sophie Rommel, Lotje de Witte, Siobhan M. Dolan, Whitney Lieb, Erona Ibroci, Sophie Ohrn, Jezelle Lynch, Christina Capuano, Daniel Stadlbauer, Florian Krammer, Lauren B. Zapata, Rachel I. Brody, Rhoda S. Sperling, Omara Afzal, Mr Roy Missall, Amy Balbierz, Teresa Janevic, Joanne Stone, Elizabeth A. Howell, Veerle Bergink

## Abstract

**Background:** In May-July 2020 in the New York City area, up to 16% of pregnant women had reportedly been infected with SARS-CoV-2. Prior studies found associations between SARS-CoV-2 infection during pregnancy and certain adverse outcomes (e.g., preterm birth, cesarean delivery). These studies relied on reverse transcription polymerase chain reaction (RT-PCR) testing to establish SARS-CoV-2 infection. This led to overrepresentation of symptomatic or acutely ill cases in scientific studies.

**Objective:** To expand our understanding of the effects of SARS-CoV-2 infection during pregnancy on pregnancy outcomes, regardless of symptomatology and stage of infection, by using serological tests to measure IgG antibody levels.

**Study Design:** The Generation C Study is an ongoing prospective cohort study conducted at the Mount Sinai Health System. All pregnant women receiving obstetrical care at the Mount Sinai Hospital and Mount Sinai West Hospital from April 20, 2020 onwards are eligible for participation. For the current analysis, we included participants who had given birth to a liveborn singleton infant on or before August 15, 2020. Blood was drawn as part of routine clinical care; for each woman, we tested the latest sample available to establish seropositivity using a SARS-CoV-2 serologic enzyme-linked immunosorbent assay. Additionally, RT-PCR testing was performed on a nasopharyngeal swab taken during labor and delivery. Pregnancy outcomes of interest (i.e., gestational age at delivery, birth weight, mode of delivery, Apgar score, ICU/NICU admission, and neonatal hospital length of stay) and covariates were extracted from electronic medical records. Among all Generation C participants who had given birth by August 15, 2020 (n=708), we established the SARS-CoV-2 seroprevalence. Excluding women who tested RT-PCR positive at delivery, we conducted crude and adjusted linear and logistic regression models to compare antibody positive women without RT-PCR positivity at delivery with antibody negative women without RT-PCR positivity at delivery. We stratified analyses by race/ethnicity to examine potential effect modification.

**Results:** The SARS-CoV-2 seroprevalence based on IgG measurement was 16.4% (n=116, 95% CI 13.7-19.3). Twelve women (1.7%) were SARS-CoV-2 RT-PCR positive at delivery (11 of these women were seropositive). Seropositive women were generally younger, more often Black or Hispanic, and more often had public insurance and higher pre-pregnancy BMI compared with seronegative women. SARS-CoV-2 seropositivity without RT-PCR positivity at delivery was associated with decreased odds of caesarean delivery (aOR 0.48, 95%CI 0.27; 0.84) compared with seronegative women without RT-PCR positivity at delivery. Stratified by race/ethnicity, the association between seropositivity and decreased odds of caesarean delivery remained for non-Hispanic Black/African-American and Hispanic women, but not for non-Hispanic White women. No other pregnancy outcomes differed by seropositivity, overall or stratified by race/ethnicity.

**Conclusion:** Seropositivity for SARS-CoV-2 without RT-PCR positivity at delivery, suggesting that infection occurred earlier during pregnancy, was not associated with selected adverse maternal or neonatal outcomes among live births in a cohort sample of women from New York City. While non-Hispanic Black and Latina women in our cohort had a higher rate of SARS-CoV-2 seropositivity compared with non-Hispanic White women, we found no increase in adverse maternal or neonatal outcomes among these groups due to infection.

## Introduction

The coronavirus disease 2019 (COVID-19) pandemic is an ongoing global health threat, caused by the severe acute respiratory syndrome coronavirus 2 (SARS-CoV-2). With 16,519,668 confirmed cases and 302,992 confirmed deaths as of December 17, 2020, the United States is currently one of the most affected countries worldwide ^1^. Despite the widespread prevalence of the virus, questions remain about how SARS-CoV-2 impacts vulnerable populations, including pregnant women. Recent findings suggest that in the New York City area, up to 16% of pregnant women have been infected with SARS-CoV-2 ^2,3^.

Efforts to research maternal and neonatal outcomes in pregnant women with confirmed SARS-CoV-2 infection are ongoing worldwide to provide the general public, medical professionals, and policy makers with crucial information about the potential consequences of SARS-CoV-2 infection during pregnancy. Although the absolute risks for severe SARS-CoV-2-related outcomes among pregnant women are low, recent research indicates that pregnant women with symptomatic SARS-CoV-2 infection have a higher mortality risk and are more likely to require intensive care unit (ICU) admission and invasive ventilation compared with age-matched nonpregnant women ^4,5^. In addition, existing studies find associations between SARS-CoV-2 infection during pregnancy and certain adverse pregnancy outcomes such as preterm birth and caesarean delivery ^2,6–8^. These studies, however, rely on reverse transcription polymerase chain reaction (RT-PCR) testing to establish SARS-CoV-2 infection. RT-PCR testing is limited; it only identifies active or very recent infections and is usually performed based on clinical indication which may lead to an overrepresentation of symptomatic cases in scientific studies. For pregnant women, RT-PCR nasopharyngeal testing may be universally performed upon admission for labor and delivery, but not earlier in pregnancy. Unless RT-PCR testing is frequently and routinely administered to all pregnant women, many infections will be missed. To understand the consequences of SARS-CoV-2 infection during pregnancy for women and their newborns, outcomes should be examined in women with and without SARS-CoV-2 infection at any point during pregnancy, regardless of their symptomatology. Another way to detect SARS-CoV-2 infection is using serological tests to measure IgG antibody levels. Although not without limitations, the advantage of serological testing is that it can identify individuals previously infected with SARS-CoV-2, even if they were asymptomatic and/or never underwent testing while acutely infected.

To this end, we started the Generation C Study, an ongoing prospective cohort study aiming to enroll 4,000 pregnant women in New York City (NYC), with assessments of SARS-CoV-2 infection using serological testing throughout pregnancy and PCR testing at delivery. We considered a seropositive sample as an indicator of SARS-CoV-2 infection during pregnancy because the first SARS-CoV-2 infection in NYC was confirmed on March 1, 2020 and our study included women who gave birth on or before August 15, 2020. Here we report interim findings from the first 708 women who participated between April and August 2020 and for whom pregnancy outcomes were available. Our objectives were to describe the SARS-CoV-2 seroprevalence among pregnant women in our cohort and to examine the associations between serostatus without RT-PCR positivity at delivery and select adverse maternal and neonatal outcomes.

## Materials and Methods

### Study design and participants

The Generation C Study is a prospective cohort study designed to examine the impact of SARS-CoV-2 infection and immune response in pregnant women (symptomatic and asymptomatic) on maternal, fetal, and neonatal outcomes. The Generation C Study utilizes existing infrastructure to collect blood samples from pregnant women throughout pregnancy to test for IgG antibodies to SARS-CoV-2. The study is being conducted at the Mount Sinai Health System (MSHS), the largest healthcare system in NYC, which has over 14,000 deliveries each year. All pregnant women receiving obstetrical care at the Mount Sinai Hospital and Mount Sinai West Hospital (two MSHS hospital campuses located in Manhattan) during the study period are eligible for participation. Recruitment and sampling started on April 20, 2020 and are currently ongoing. Given that women are seen multiple times during pregnancy and blood is drawn during routine prenatal care, some women have more than one blood sample available. For the current interim analysis, we focused on participants who gave birth to a liveborn singleton infant on or before August 15, 2020; we excluded (from this interim analysis) participants with other outcomes (e.g., miscarriage, abortion, stillbirth) due to limited statistical power (n=3). For each woman, we selected the latest sample available (either collected during a second or third trimester prenatal visit or upon admission to labor and delivery) to establish serostatus. Women were informed about the study before their obstetrical care appointment through printed materials, emails, and information posted in online patient portals. All participants provided informed consent per the institutional review board (IRB)-approved study protocol (IRB at the Icahn School of Medicine at Mount Sinai, protocol IRB-20-03352, April 15, 2020).

### Serology testing

The Generation C Study employed a serologic enzyme-linked immunosorbent assay (ELISA) developed at the Icahn School of Medicine at Mount Sinai ^9^. This assay is based on the soluble receptor-binding domain and the trimerized, stabilized full-length spike protein. The assay used in this study closely resembles an assay established in the MSHS CLIA-certified Clinical Pathology Laboratory, which received New York State Department of Health (NYSDOH) and Food and Drug Administration (FDA) emergency use authorization (EUA) in early 2020 ^9,10^. The test has high sensitivity (95.0%) and specificity (100%), as determined with an initial validation panel of samples, with a positive predictive value of 100%, and a negative predictive value of 97.0% ^11^. We measured IgG antibodies because this type of antibody is produced for at least three months and potentially longer after exposure ^12–14^. Whereas some studies with small numbers of participants have shown rapid decay of SARS-CoV-2 antibodies over time ^15,16^, a recent examination of the assay being used in the Generation C study found that the vast majority of infected individuals with mild-to moderate COVID-19 experienced robust IgG antibody responses against the viral spike protein and that the titers were relatively stable for at least five months ^17^ (cite). By using a low dilution (1:50) for the screening assay, we tested for SARS-CoV-2 seropositivity. Positive samples were further diluted and tested in an assay using the full-length spike protein to determine the antibody endpoint titer.

### Molecular testing

Beginning March 27, 2020, MSHS implemented universal molecular testing for all pregnant women admitted for labor and delivery. A nucleic acid RT-PCR test to detect SARS-CoV-2 is performed on a nasopharyngeal RT-PCR swab sample obtained at the time of labor and delivery admission.

### Pregnancy outcomes and covariates

Pregnancy outcomes of interest and covariates were extracted from the electronic medical records (EMR) of participating women. Outcomes examined were gestational age at delivery, birth weight, mode of delivery, Apgar score at 5 minutes, maternal ICU admission, neonatal ICU (NICU) admission, neonatal hospital length of stay, and maternal and neonatal mortality during hospitalization (or known follow-up up to six months among those continuing care at MSHS). The analyses were adjusted in a step-wise procedure for the following covariates, which are potential risk factors for both SARS-CoV-2 seropositivity or infection severity ^18–23^ and adverse pregnancy outcomes ^24–29^: maternal age, parity, race/ethnicity, insurance status, tobacco use during pregnancy, alcohol use during pregnancy, illicit drug use during pregnancy (e.g., marijuana, cocaine), pre-pregnancy body mass index (BMI), pre-pregnancy diabetes, and pre-pregnancy hypertension.

### Statistical analysis

First, we calculated the SARS-CoV-2 seroprevalence in our sample as the number of women with SARS-CoV-2 spike IgG antibodies divided by the total number of women in the sample and constructed a 95% confidence interval around the estimate. We also estimated the proportion of women testing RT-PCR positive at delivery. Of note, 7.6% of women in our cohort had missing RT-PCR test result data at delivery and were assumed to be negative. Given the small number of women testing RT-PCR positive at delivery (n=12) and the likelihood that these women received differential care potentially resulting in altered outcomes, these women were excluded from analyses of associations between serostatus and selected adverse pregnancy outcomes; we hope to include these women in future analyses. We then categorized women into one of two groups: 1) antibody negative without RT-PCR positivity at delivery [reference group]); or 2) antibody positive without RT-PCR positivity at delivery.

Differences between seronegative women without RT-PCR positivity at delivery and seropositive women without RT-PCR positivity at delivery were examined using the appropriate tests, including Fisher’s exact test, chi-square test, or t-test. To examine the effect of SARS-CoV-2 seropositivity during pregnancy on outcomes of interest among live births, we conducted crude and adjusted linear and logistic regression models (depending on the nature of the outcome variable) to compare seropositive women with seronegative women. To account for potential effect modification, we additionally stratified models by race/ethnicity. We performed sensitivity analyses excluding participants with a missing RT-PCR result at delivery. In a second series of sensitivity analyses, we excluded those participants for whom the time between their latest collected blood sample and delivery was more than 30 days to avoid misclassification of women with SARS-CoV-2 infection later in pregnancy as seronegative. SPSS software version 26.0 was used for data analysis.

## Results

A total of 708 Generation C participants had given birth by August 15, 2020. Mean gestational age at time of serosample collection was 259.2 days (SD 27.3) and mean time between serosample collection and delivery was 13.5 days (SD 24.7). Most serosamples (n=448, 63.3%) were taken upon admission to labor and delivery, 255 serosamples (36.0%) were taken during a prenatal visit in the third trimester, and only five serosamples (0.7%) were taken during a prenatal visit in the second trimester. The overall SARS-CoV-2 seroprevalence based on IgG measurement (regardless of SARS-CoV-2 RT-PCR test result at delivery) was 16.4% (n=116, 95% confidence interval [CI] 13.7-19.3). Among these seropositive women, only four (3.4%) had a positive SARS-CoV-2 RT-PCR test during pregnancy, performed as part of clinical care, potentially indicating symptomatic disease. Additionally, twelve women (1.7%) were SARS-CoV-2 RT-PCR positive at delivery (11 of these women were also seropositive). Sample characteristics for seronegative and seropositive women, excluding those women with RT-PCR positivity at delivery, are shown in Table 1. Seronegative and seropositive women without RT-PCR positivity at delivery differed by maternal age, race/ethnicity, insurance status, and pre-pregnancy BMI. Seropositive women without RT-PCR positivity at delivery were generally younger, more often non-Hispanic Black/African-American or Hispanic, and more often had public insurance and higher pre-pregnancy BMI compared with seronegative women without RT-PCR positivity at delivery.

**Table 1.** Characteristics of women delivering a singleton infant according to SARS-CoV-2 IgG antibody status^*^, Mount Sinai Health System, April 20, 2020-August 15, 2020

| Characteristic | SARS-CoV-2 IgG antibody | SARS-CoV-2 IgG antibody | p-value |
| --- | --- | --- | --- |
|  | negative, n=591 | positive, n=105 |  |
| Maternal age in years, mean (SD) | 33.3 (5.2) | 31.8 (5.9) | <0.01 |
| Nulliparous, n (%) | 306 (51.8) | 44 (41.9) | 0.07 |
| Previous caesarean section, n (%) | 83 (14.0) | 13 (12.4) | 0.76 |
| Race/Ethnicity, n (%) |  |  |  |
| White, non-Hispanic | 267 (45.2) | 26 (24.8) | <0.01 |
| Black/African-American, non- | 85 (14.4) | 25 (23.8) |  |
| Hispanic | 133 (22.5) | 45 (42.9) |  |
| Hispanic | 72 (12.2) | 4 (3.8) |  |
| Asian | 25 (4.2) | 3 (2.9) |  |
| Other | 9 (1.5) | 2 (1.9) |  |
| Unknown |  |  |  |
| Insurance, n (%) |  |  |  |
| Private | 449 (76.0) | 59 (56.2) | <0.01 |
| Public | 133 (22.5) | 43 (41.0) |  |
| Self-pay | 9 (1.5) | 3 (2.9) |  |
| Tobacco use during pregnancy, n (%) | 29 (4.9) | 2 (1.9) | 0.21 |
| Alcohol use during pregnancy, n (%) | 176 (29.8) | 25 (23.8) | 0.24 |
| Illicit drug use during pregnancy, n (%) | 30 (5.1) | 5 (4.8) | 1.0 |
| Pre-pregnancy BMI, median (range) | 25.2 (16.6-59.7) | 28.0 (18.1-45.2) | <0.01 |
| Pre-pregnancy diabetes, n (%) | 4 (0.7) | 1 (1.0) | 0.56 |
| Pre-pregnancy hypertension, n (%) | 13 (2.2) | 1 (1.0) | 0.71 |

Antibody results based on testing latest available blood samples. Percentages shown are column percentages.
\*Excluding women with RT-PCR positivity, as tested using a nasopharyngeal swab at time of delivery (n=12).

### Pregnancy outcomes

Pregnancy outcomes for seropositive and seronegative women without RT-PCR positivity at delivery are summarized in Table 2. Most delivery outcomes did not significantly differ between groups, before or after adjustment (Table 3). We observed no maternal or neonatal mortality while in care at the MSHS. Only one maternal ICU admission occurred after birth, in a woman who was SARS-CoV-2 seronegative without RT-PCR positivity.

**Table 2.** Neonatal outcomes of women delivering a singleton infant according to SARS-CoV-2 IgG antibody status^*^, Mount Sinai System, April 20, 2020-August 15, 2020

| Outcome | SARS-CoV-2 IgG antibody | SARS-CoV-2 IgG antibody |
| --- | --- | --- |
|  | negative, n=591 | positive, n=105 |
| Gestational age in days, mean (SD) | 273.5 (10.4) | 271.7 (14.5) |
| Preterm birth (<37 weeks), n (%) | 37 (6.3) | 8 (7.6) |
| Birth weight in grams, mean (SD) | 3,291.3 (500.6) | 3,212.5 (538.9) |
| Small for gestational age, n (%) | 43 (7.3) | 9 (8.6) |
| Large for gestational age, n (%) | 14 (2.4) | 2 (1.9) |
| Delivery mode, n (%) |  |  |
| Spontaneous vaginal delivery | 370 (62.6) | 76 (72.4) |
| Assisted vaginal delivery | 30 (5.1) | 7 (6.7) |
| Caesarean delivery | 191 (32.3) | 22 (21.0) |
| Apgar score 5 minutes, mean (SD) | 8.9 (0.4) | 8.8 (0.9) |
| NICU admission, n (%) | 53 (9.0) | 11 (10.5) |
| Neonatal hospital length of stay in days, mean (SD) | 2.4 (3.7) | 2.4 (4.1) |

\*Excluding women with RT-PCR positivity, as tested using a nasopharyngeal swab at time of delivery (n=12).

**Table 3.**
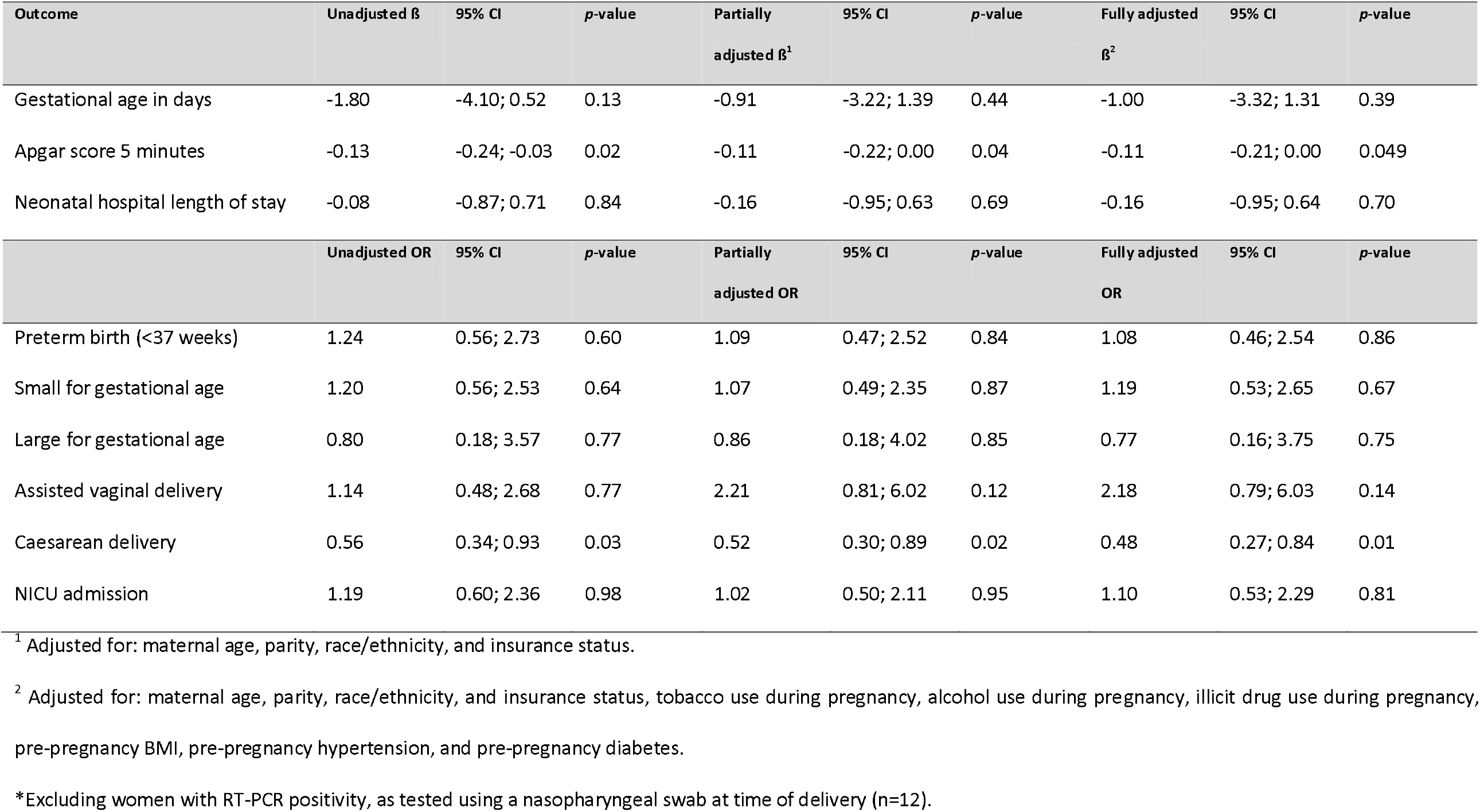
Linear and logistic regression analyses of delivery and neonatal outcomes comparing SARS-CoV-2 IgG antibody positive women with IgG antibody negative women^*^

| Outcome | Unadjusted $\beta$ | 95% CI | p-value | Partially<br>adjusted $\beta^1$ | 95% CI | p-value | Fully adjusted<br>$\beta^2$ | 95% CI | p-value |
| --- | --- | --- | --- | --- | --- | --- | --- | --- | --- |
| Gestational age in days | -1.80 | -4.10; 0.52 | 0.13 | -0.91 | -3.22; 1.39 | 0.44 | -1.00 | -3.32; 1.31 | 0.39 |
| Apgar score 5 minutes | -0.13 | -0.24; -0.03 | 0.02 | -0.11 | -0.22; 0.00 | 0.04 | -0.11 | -0.21; 0.00 | 0.049 |
| Neonatal hospital length of stay | -0.08 | -0.87; 0.71 | 0.84 | -0.16 | -0.95; 0.63 | 0.69 | -0.16 | -0.95; 0.64 | 0.70 |
|  | Unadjusted OR | 95% CI | p-value | Partially<br>adjusted OR | 95% CI | p-value | Fully adjusted<br>OR | 95% CI | p-value |
| Preterm birth (<37 weeks) | 1.24 | 0.56; 2.73 | 0.60 | 1.09 | 0.47; 2.52 | 0.84 | 1.08 | 0.46; 2.54 | 0.86 |
| Small for gestational age | 1.20 | 0.56; 2.53 | 0.64 | 1.07 | 0.49; 2.35 | 0.87 | 1.19 | 0.53; 2.65 | 0.67 |
| Large for gestational age | 0.80 | 0.18; 3.57 | 0.77 | 0.86 | 0.18; 4.02 | 0.85 | 0.77 | 0.16; 3.75 | 0.75 |
| Assisted vaginal delivery | 1.14 | 0.48; 2.68 | 0.77 | 2.21 | 0.81; 6.02 | 0.12 | 2.18 | 0.79; 6.03 | 0.14 |
| Caesarean delivery | 0.56 | 0.34; 0.93 | 0.03 | 0.52 | 0.30; 0.89 | 0.02 | 0.48 | 0.27; 0.84 | 0.01 |
| NICU admission | 1.19 | 0.60; 2.36 | 0.98 | 1.02 | 0.50; 2.11 | 0.95 | 1.10 | 0.53; 2.29 | 0.81 |

<sup>1</sup> Adjusted for: maternal age, parity, race/ethnicity, and insurance status.
<sup>2</sup> Adjusted for: maternal age, parity, race/ethnicity, and insurance status, tobacco use during pregnancy, alcohol use during pregnancy, illicit drug use during pregnancy, pre-pregnancy BMI, pre-pregnancy hypertension, and pre-pregnancy diabetes.
\*Excluding women with RT-PCR positivity, as tested using a nasopharyngeal swab at time of delivery (n=12).

There was a significant difference in the prevalence of caesarean delivery (CD) between the two groups (32.3% in seronegative women without RT-PCR positivity at delivery compared with 21.0% in seropositive women without RT-PCR positivity at delivery). After adjustment, seropositivity without RT-PCR positivity at delivery was associated with decreased odds of CD (fully adjusted odds ratio [aOR] 0.48, 95%CI 0.27; 0.84). When stratified by race/ethnicity, SARS-CoV-2 seropositivity without RT-PCR positivity at delivery was associated with decreased odds of CD in non-Hispanic Black/African-American and Hispanic women (aOR 0.23, 95%CI 0.07; 0.78 and aOR 0.27, 95%CI 0.10; 0.71, respectively), but not in non-Hispanic White women (aOR 1.19, 95%CI 0.46; 3.10, data not shown).

Additionally, seropositive women without RT-PCR positivity at delivery had slightly lower Apgar scores at 5 minutes (adjusted *ß* −0.11, 95%CI −0.21; 0.00), of which the clinical relevance is limited given the overall high Apgar scores. Stratified by race/ethnicity, results were no longer significant. Associations between seropositivity and the other outcome variables did not vary by race/ethnicity.

The sensitivity analyses, which excluded (1) participants with a missing RT-PCR result at delivery, and (2) participants with more than 30 days between serosample collection and delivery, produced similar results (Supplemental Tables 1 and 2).

## Discussion

### Principal findings

This preliminary analysis of participants from a prospective cohort study of pregnant women shows that seropositivity for SARS-CoV-2 in the absence of detected infection at delivery using a nasopharyngeal RT-PCR (suggesting that infection occurred earlier during pregnancy) was not associated with selected adverse maternal or neonatal outcomes among live births in our sample of women from NYC.

### Results

These findings contrast with findings from systematic reviews that found SARS-CoV-2 infection to be associated with increased risks of preterm birth ^6,7,30^. However, most previous research used a single RT-PCR test to confirm SARS-CoV-2 infection, when indicated or as part of universal screening at time of delivery. Symptomatic women and women with active infection at delivery might be over-represented in these studies, which might be missing women with resolved infections or ongoing infections that are no longer positive by nasopharyngeal RT-PCR testing. By measuring SARS-CoV-2 IgG antibodies, we were able to study SARS-CoV-2 exposure earlier in pregnancy irrespective of symptomatology and testing while acutely infected.

Similar to previous work in both the general and pregnant populations ^31–34^, we show that SARS-CoV-2 disproportionately affects groups that have been economically/socially marginalized. Black and Hispanic women, as well as women with public insurance, had higher proportions of SARS-CoV-2 seropositivity compared with non-Hispanic White women and women with private insurance. These findings may be explained by various factors disproportionally impacting Black and Hispanic women and women with a lower socioeconomic status (SES), such as segregated neighborhoods, crowded housing, discrimination or a woman’s occupation as an essential worker ^35,36^. In addition to their increased risk of contracting SARS-CoV-2, research has also documented disparities in maternal and neonatal outcomes by race/ethnicity and SES ^37–39^. In contrast, our findings do not indicate that SARS-CoV-2 infection adds to their elevated risk of adverse maternal and neonatal outcomes. However, these preliminary findings require confirmation in a larger sample.

Seropositivity in our sample was associated with a decreased risk of CD. In contrast, previous research showed that SARS-CoV-2 infection was associated with a higher risk of CD, with the risk for CD increasing with increasing severity of maternal illness ^40^. As previously mentioned, in many of these studies, symptomatic women and women with an active or ongoing infection at delivery are overrepresented. It is conceivable that in (severely) ill mothers, clinicians are more likely to perform a CD to avoid adverse perinatal and neonatal outcomes. Our seropositive group consisted of women who contracted SARS-CoV-2 before going into labor, and who had a negative RT-PCR test at time of delivery. One other study, in which SARS-CoV-2 infection during pregnancy was evaluated using antibody testing in a Danish sample, did not find a difference in CD rate between antibody positive and negative women. However, the absolute number of seropositive women in that study was low (n=28) ^41^. Our findings may be influenced by the small sample size, variation in clinical practice, or unmeasured confounding factors. If a decrease in CD rate in seropositive women without RT-PCR positivity at delivery is confirmed by further research, future studies should explore the underlying causal pathways which are unaccounted for in the current analysis.

We found that the active infection rate based on RT-PCR at delivery in our sample was similar to the MSHS-wide infection rate based on RT-PCR at delivery in the corresponding months, suggesting that our sample is representative of the MSHS pregnant population. However, due to the small number (n=12) of women who tested positive for SARS-CoV-2 on RT-PCR testing at delivery in this interim analysis, we chose not to examine them separately in analyses of associations with pregnancy outcomes. We will include these women in a future analysis of the full sample (target N=4,000).

### Clinical implications

We found no indication of adverse maternal or neonatal outcomes among live births related to SARS-CoV-2 seropositivity detected during pregnancy among women from NYC. Our findings, therefore, provide some reassurance regarding the effects of SARS-CoV-2 infection during pregnancy. However, since these findings are based on a subsample of our cohort and a selection of key maternal and neonatal outcomes, further research is needed to strengthen the evidence base on the effects of SARS-CoV-2 infection during pregnancy on pregnancy outcomes.

### Research implications

Research indicates that inflammatory responses earlier in pregnancy might produce more marked adverse effects on the fetus (e.g. on neurodevelopment) than those that occur later ^42–44^. Future analyses might measure seropositivity within each trimester of pregnancy to better pinpoint when each woman became infected and how this timing may impact the outcome of her pregnancy and the health of her baby.

### Strengths and limitations

We measured antibodies with a very sensitive test right after the start of the pandemic in an ethnically and socially diverse sample of pregnant women. The findings of this analysis are subject to limitations. First, we measured SARS-CoV-2 seropositivity in the second and third trimester during antenatal care or at labor and delivery. Consequently, we cannot be certain when these women were infected with SARS-CoV-2. However, since widespread community transmission of SARS-CoV-2 began in March 2020, we can be sure that the infection occurred during pregnancy since all women included in this study delivered by mid-August 2020. Second, due to the putative decay of SARS-CoV-2 antibodies in milder COVID-19 cases over time ^16^, we cannot preclude potential misclassification of women as seronegative who were infected earlier in pregnancy and no longer produced antibodies at the time of blood sampling. Although our study was designed to collect multiple blood samples from participants during each trimester of pregnancy, very few participants in the current analysis had repeat blood samples; this precluded us from examining potential seroconversion throughout pregnancy. However, recent findings about the serologic assay used in our study indicate that robust antibodies to SARS-CoV-2 infection persist for at least five months in the majority of the people ^17^. Third, we were unable to obtain information on presence of symptoms which may impact maternal and infant outcomes. However, many studies to date have only focused on women with symptomatic SARS-CoV-2 infection. We consider it a strength to be able to look at the effects of both symptomatic and asymptomatic SARS-CoV-2 infection. Fourth, our sample of women testing RT-PCR positive for SARS-CoV-2 infection at time of delivery was small, precluding us from examining this group of women separately. However, data collection is ongoing, and we expect to have a larger sample at the end of the study. Fifth, we only included liveborn infants in our current analysis due to small numbers of other outcomes; we intend to report on the prevalence of miscarriage and stillbirths at the end of the study. Last, although our cohort included a diverse sample of pregnant women, we were unable to assess the representativeness of our cohort since recruitment procedures precluded our ability to determine the proportion of women eligible who enrolled or compare characteristics of women eligible who enrolled versus did not enroll.

## Conclusion

Seropositivity for SARS-CoV-2 without RT-PCR positivity at delivery, suggesting that infection occurred earlier during pregnancy, was not associated with increased odds of selected adverse maternal or neonatal outcomes among live births in a cohort sample of women from the Mount Sinai Health System, NYC (April-August 2020). While non-Hispanic Black/African-American and Latina women were more likely to contract the virus, we found no increase in adverse maternal or neonatal outcomes among these groups due to infection.

## Data Availability

Data not publicly available. Please contact the corresponding author for data requests.

## Acknowledgements

The authors would like to thank several members of the US Centers for Disease Control (CDC) that have contributed to the interpretation of the data and have provided their feedback on the manuscript: Margaret C. Snead, Sascha R. Ellington, Romeo R. Galang, Suzanne M. Gilboa, Kate R. Woodworth, Titilope Oduyebo, Ashley Smoots, and Laura D. Zambrano. The authors also thank Dr. Michael Brodman, Dr. Alan Adler and Dr. Francesco Callipari in the Department of Obstetrics, Gynecology and Reproductive Science at the Mount Sinai Health System for their efforts to optimize participant recruitment. They did not receive any financial compensation for their contribution.

---

**Supplementary Table 1.** Sensitivity analyses of linear and logistic regression analyses of delivery and neonatal outcomes comparing SARS-CoV-2 IgG antibody positive women with IgG antibody negative women^*^, excluding women with a missing RT-PCR test at delivery

| Outcome | Unadjusted $\beta$ | 95% CI | p-value | Partially<br>adjusted $\beta^1$ | 95% CI | p-value | Fully adjusted<br>$\beta^2$ | 95% CI | p-value |
| --- | --- | --- | --- | --- | --- | --- | --- | --- | --- |
| Gestational age in days | -1.57 | -4.07; 0.92 | 0.22 | -0.78 | -3.27; 1.73 | 0.54 | -0.76 | -3.27; 1.74 | 0.55 |
| Apgar score 5 minutes | -0.15 | -0.27; -0.04 | 0.01 | -0.14 | -0.26; -0.02 | 0.02 | -0.13 | -0.24; -0.01 | 0.03 |
| Neonatal hospital length of stay | -0.02 | -0.89; 0.85 | 0.96 | -0.09 | -0.96; 0.79 | 0.85 | -0.11 | -0.98; 0.76 | 0.81 |
|  | Unadjusted OR | 95% CI | p-value | Partially<br>adjusted OR | 95% CI | p-value | Fully adjusted<br>OR | 95% CI | p-value |
| Preterm birth (<37 weeks) | 1.32 | 0.59; 2.93 | 0.50 | 1.19 | 0.51; 2.78 | 0.68 | 1.18 | 0.50; 2.79 | 0.71 |
| Small for gestational age | 1.02 | 0.44; 2.35 | 0.96 | 0.87 | 0.36; 2.06 | 0.74 | 0.95 | 0.40; 2.35 | 0.95 |
| Large for gestational age | 1.00 | 0.22; 4.53 | 1.00 | 1.03 | 0.21; 5.00 | 0.97 | 1.00 | 0.20; 5.09 | 1.00 |
| Assisted vaginal delivery | 1.17 | 0.46; 2.95 | 0.74 | 2.11 | 0.73; 6.14 | 0.17 | 2.07 | 0.69; 6.15 | 0.19 |
| Caesarean delivery | 0.51 | 0.29; 0.88 | 0.02 | 0.46 | 0.25; 0.83 | 0.01 | 0.41 | 0.22; 0.76 | <0.01 |
| NICU admission | 1.08 | 0.51; 2.29 | 0.83 | 0.92 | 0.42; 2.02 | 0.83 | 0.96 | 0.43; 2.15 | 0.93 |

<sup>1</sup> Adjusted for: maternal age, parity, race/ethnicity, and insurance status.
<sup>2</sup> Adjusted for: maternal age, parity, race/ethnicity, and insurance status, tobacco use during pregnancy, alcohol use during pregnancy, illicit drug use during pregnancy, pre-pregnancy BMI, pre-pregnancy hypertension, and pre-pregnancy diabetes.
\*Excluding women with RT-PCR positivity, as tested using a nasopharyngeal swab at time of delivery (n=12).

**Supplementary Table 2.**
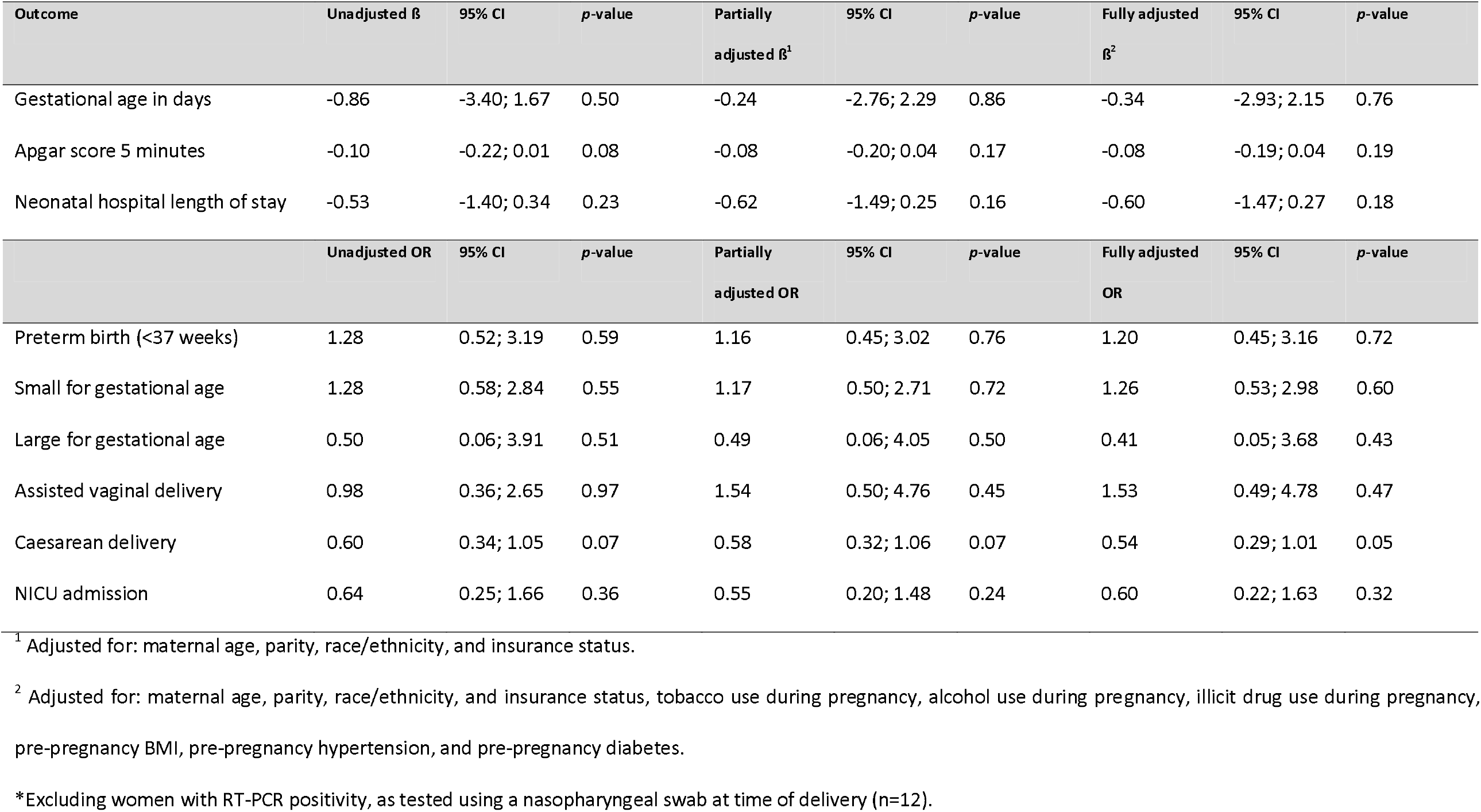
Sensitivity analyses of linear and logistic regression analyses of delivery and neonatal outcomes comparing SARS-CoV-2 IgG antibody positive women with IgG antibody negative women^*^, excluding women ≥30 days between sample collection and delivery

| Outcome | Unadjusted $\beta$ | 95% CI | p-value | Partially adjusted $\beta^1$ | 95% CI | p-value | Fully adjusted $\beta^2$ | 95% CI | p-value |
| --- | --- | --- | --- | --- | --- | --- | --- | --- | --- |
| Gestational age in days | -0.86 | -3.40; 1.67 | 0.50 | -0.24 | -2.76; 2.29 | 0.86 | -0.34 | -2.93; 2.15 | 0.76 |
| Apgar score 5 minutes | -0.10 | -0.22; 0.01 | 0.08 | -0.08 | -0.20; 0.04 | 0.17 | -0.08 | -0.19; 0.04 | 0.19 |
| Neonatal hospital length of stay | -0.53 | -1.40; 0.34 | 0.23 | -0.62 | -1.49; 0.25 | 0.16 | -0.60 | -1.47; 0.27 | 0.18 |
|  | Unadjusted OR | 95% CI | p-value | Partially adjusted OR | 95% CI | p-value | Fully adjusted OR | 95% CI | p-value |
| Preterm birth (<37 weeks) | 1.28 | 0.52; 3.19 | 0.59 | 1.16 | 0.45; 3.02 | 0.76 | 1.20 | 0.45; 3.16 | 0.72 |
| Small for gestational age | 1.28 | 0.58; 2.84 | 0.55 | 1.17 | 0.50; 2.71 | 0.72 | 1.26 | 0.53; 2.98 | 0.60 |
| Large for gestational age | 0.50 | 0.06; 3.91 | 0.51 | 0.49 | 0.06; 4.05 | 0.50 | 0.41 | 0.05; 3.68 | 0.43 |
| Assisted vaginal delivery | 0.98 | 0.36; 2.65 | 0.97 | 1.54 | 0.50; 4.76 | 0.45 | 1.53 | 0.49; 4.78 | 0.47 |
| Caesarean delivery | 0.60 | 0.34; 1.05 | 0.07 | 0.58 | 0.32; 1.06 | 0.07 | 0.54 | 0.29; 1.01 | 0.05 |
| NICU admission | 0.64 | 0.25; 1.66 | 0.36 | 0.55 | 0.20; 1.48 | 0.24 | 0.60 | 0.22; 1.63 | 0.32 |

<sup>1</sup> Adjusted for: maternal age, parity, race/ethnicity, and insurance status.
<sup>2</sup> Adjusted for: maternal age, parity, race/ethnicity, and insurance status, tobacco use during pregnancy, alcohol use during pregnancy, illicit drug use during pregnancy, pre-pregnancy BMI, pre-pregnancy hypertension, and pre-pregnancy diabetes.
\*Excluding women with RT-PCR positivity, as tested using a nasopharyngeal swab at time of delivery (n=12).

## Notes

**Role of funding source** This study is partially funded (contract 75D30120C08186) by the US Centers for Disease Control and Prevention (CDC), who also provided technical assistance related to analysis and interpretation of data and writing the report. The findings and conclusions in this report are those of the authors and do not necessarily represent the official position of the CDC. Initial assay development work in the Krammer laboratory was partially supported by the NIAID Centers of Excellence for Influenza Research and Surveillance (CEIRS) contract HHSN272201400008C (FK, for reagent generation), Collaborative Influenza Vaccine Innovation Centers (CIVIC) contract 75N93019C00051 (FK, for reagent generation), and the generous support of the JPB foundation, the Open Philanthropy Project (#2020-215611) and other philanthropic donations. These funding sources were not involved in the current study.

### Competing Interest Statement

Mount Sinai has licensed serological assays to commercial entities and has filed for patent protection for serological assays. D.S and F.K. are listed as inventors on the pending patent application. The other authors have nothing to report.

### Funding Statement

This study is partially funded (contract 75D30120C08186) by the US Centers for Disease Control and Prevention (CDC), who also provided technical assistance related to analysis and interpretation of data and writing the report. The findings and conclusions in this report are those of the authors and do not necessarily represent the official position of the CDC.
Initial assay development work in the Krammer laboratory was partially supported by the NIAID Centers of Excellence for Influenza Research and Surveillance (CEIRS) contract HHSN272201400008C (FK, for reagent generation), Collaborative Influenza Vaccine Innovation Centers (CIVIC) contract 75N93019C00051 (FK, for reagent generation), and the generous support of the JPB foundation, the Open Philanthropy Project (#2020-215611) and other philanthropic donations. These funding sources were not involved in the current study.

### Author Declarations

All participants provided informed consent per the institutional review board (IRB)-approved study protocol (IRB at the Icahn School of Medicine at Mount Sinai, protocol IRB-20-03352, April 15, 2020).

## References

1. CDC. CDC COVID Data Tracker. https://covid.cdc.gov/covid-data-tracker/?CDC_AA_refVal=https%3A%2F%2Fwww.cdc.gov%2Fcoronavirus%2F2019-ncov%2Fcases-updates%2Fcases-in-us.html#cases_casesper100klast7days. Accessed March 12, 2020.

2. Prabhu M, Cagino K, Matthews KC, et al. Pregnancy and postpartum outcomes in a universally tested population for SARS-CoV-2 in New York City: A prospective cohort study. BJOG. July 2020. doi:10.1111/1471-0528.16403

3. Haizler-Cohen L, Davidov A, Blitz MJ, Fruhman G. Severe acute respiratory syndrome coronavirus 2 antibodies in pregnant women admitted to labor and delivery units. Am J Obstet Gynecol. 2020. doi:10.1016/j.ajog.2020.09.022

4. Zambrano LD, Ellington S, Strid P, et al. Update: Characteristics of Symptomatic Women of Reproductive Age with Laboratory-Confirmed SARS-CoV-2 Infection by Pregnancy Status — United States, January 22–October 3, 2020. MMWR Morb Mortal Wkly Rep. 2020;69(44):1641–1647. doi:10.15585/mmwr.mm6944e3

5. DeBolt C, Bianco A, Limaye M, et al. Pregnant women with severe or critical COVID-19 have increased composite morbidity compared to non-pregnant matched controls. Am J Obstet Gynecol. 2020. doi:https://doi.org/10.1016/j.ajog.2020.11.022

6. Khalil A, Kalafat E, Benlioglu C, et al. SARS-CoV-2 infection in pregnancy: A systematic review and meta-analysis of clinical features and pregnancy outcomes. EClinicalMedicine. 2020;0(0):100446. doi:10.1016/j.eclinm.2020.100446

7. Allotey J, Stallings E, Bonet M, et al. Clinical manifestations, risk factors, and maternal and perinatal outcomes of coronavirus disease 2019 in pregnancy: living systematic review and meta-analysis. BMJ. 2020;370:m3320. doi:10.1136/bmj.m3320

8. Woodworth KR, Olsen E, Neelam V, Al. E. Birth and Infant Outcomes Following Laboratory-Confirmed SARS-CoV-2 Infection in Pregnancy — SET-NET, 16 Jurisdictions, March 29–October 14, 2020.; 2020. doi:http://dx.doi.org/10.15585/mmwr.mm6944e2externalicon

9. Amanat F, Stadlbauer D, Strohmeier S, et al. A serological assay to detect SARS-CoV-2 seroconversion in humans. Nat Med. 2020;26(7):1033–1036. doi:10.1038/s41591-020-0913-5

10. Stadlbauer D, Amanat F, Chromikova V, et al. SARS-CoV-2 Seroconversion in Humans: A Detailed Protocol for a Serological Assay, Antigen Production, and Test Setup. Curr Protoc Microbiol. 2020;57(1). doi:10.1002/cpmc.100

11. Stadlbauer D, Tan J, Jiang K, et al. Repeated cross-sectional sero-monitoring of SARS-CoV-2 in New York City. Nature. November 2020. doi:10.1038/s41586-020-2912-6

12. Bettencourt P, Fernandes C, Gil A, Almeida A, Alvelos M. Qualitative serology in patients recovered from SARS CoV 2 infection. Int J Pharm. 2020;81(2). doi:10.1016/j.jinf.2020.05.057

13. Wang Y, Zhang L, Sang L, et al. Kinetics of viral load and antibody response in relation to COVID-19 severity. J Clin Invest. July 2020. doi:10.1172/JCI138759

14. Seow J, Graham C, Merrick B, et al. Longitudinal evaluation and decline of antibody responses in SARS-CoV-2 infection. medRxiv. July 2020:2020.07.09.20148429. doi:10.1101/2020.07.09.20148429

15. Self WH, Tenforde MW, Stubblefield WB, et al. Decline in SARS-CoV-2 Antibodies After Mild Infection Among Frontline Health Care Personnel in a Multistate Hospital Network — 12 States, April–August 2020. MMWR Morb Mortal Wkly Rep. 2020;69(47):1762–1766. doi:10.15585/mmwr.mm6947a2

16. Ibarrondo FJ, Fulcher JA, Goodman-Meza D, et al. Rapid Decay of Anti–SARS-CoV-2 Antibodies in Persons with Mild Covid-19. N Engl J Med. 2020;383(11):1085–1087. doi:10.1056/nejmc2025179

17. Wajnberg A, Amanat F, Firpo A, et al. Robust neutralizing antibodies to SARS-CoV-2 infection persist for months. Science. 2020;370(6521). doi:10.1126/SCIENCE.ABD7728

18. Zheng Z, Peng F, Xu B, et al. Risk factors of critical & mortal COVID-19 cases: A systematic literature review and meta-analysis. J Infect. 2020;81(2):e16–e25. doi:10.1016/j.jinf.2020.04.021

19. Zhang J, Wang X, Jia X, et al. Risk factors for disease severity, unimprovement, and mortality in COVID-19 patients in Wuhan, China. Clin Microbiol Infect. 2020;26(6):767–772. doi:10.1016/j.cmi.2020.04.012

20. Jordan RE, Adab P, Cheng KK. Covid-19: Risk factors for severe disease and death. BMJ. 2020;368. doi:10.1136/bmj.m1198

21. Rod JE, Oviedo-Trespalacios O, Cortes-Ramirez J. A brief-review of the risk factors for covid-19 severity. Rev Saude Publica. 2020;54. doi:10.11606/S1518-8787.2020054002481

22. Finer N, Garnett SP, Bruun JM. COVID-19 and obesity. Clin Obes. 2020;10(3). doi:10.1111/cob.12365

23. Hamer M, Kivimäki M, Gale CR, Batty GD. Lifestyle risk factors, inflammatory mechanisms, and COVID-19 hospitalization: A community-based cohort study of 387,109 adults in UK. Brain Behav Immun. 2020;87:184–187. doi:10.1016/j.bbi.2020.05.059

24. Jaddoe VWV, Troe EJWM, Hofman A, et al. Active and passive maternal smoking during pregnancy and the risks of low birthweight and preterm birth: The generation R study. Paediatr Perinat Epidemiol. 2008;22(2):162–171. doi:10.1111/j.1365-3016.2007.00916.x

25. Catalano PM, Shankar K. Obesity and pregnancy: Mechanisms of short term and long term adverse consequences for mother and child. BMJ. 2017;356. doi:10.1136/bmj.j1

26. Macintosh MCM, Fleming KM, Bailey JA, et al. Perinatal mortality and congenital anomalies in babies of women with type 1 or type 2 diabetes in England, Wales, and Northern Ireland: Population based study. Br Med J. 2006;333(7560):177–180. doi:10.1136/bmj.38856.692986.AE

27. Maselli DJ, Adams SG, Peters JI, Levine SM. Management of asthma during pregnancy. Ther Adv Respir Dis. 2013;7(2):87–100. doi:10.1177/1753465812464287

28. Lean SC, Derricott H, Jones RL, Heazell AEP. Advanced maternal age and adverse pregnancy outcomes: A systematic review and meta-analysis. PLoS One. 2017;12(10). doi:10.1371/journal.pone.0186287

29. Adam K. Pregnancy in Women with Cardiovascular Diseases. Methodist Debakey Cardiovasc J. 2017;13(4):209–215. doi:10.14797/mdcj-13-4-209

30. Han Y, Ma H, Suo M, et al. Clinical manifestation, outcomes in pregnant women with COVID-19 and the possibility of vertical transmission: a systematic review of the current data. J Perinat Med. October 2020. doi:10.1515/jpm-2020-0431

31. COVID-19 Cases in New York City, a Neighborhood-Level Analysis – NYU Furman Center.

32. Price-Haywood EG, Burton J, Fort D, Seoane L. Hospitalization and Mortality among Black Patients and White Patients with Covid-19. N Engl J Med. 2020;382(26):2534–2543. doi:10.1056/NEJMsa2011686

33. Grechukhina O, Greenberg V, Lundsberg LS, et al. Coronavirus disease 2019 pregnancy outcomes in a racially and ethnically diverse population. Am J Obstet Gynecol MFM. October 2020:100246. doi:10.1016/j.ajogmf.2020.100246

34. Dd F, S G, MB D, et al. SARS-CoV-2 Seroprevalence Among Parturient Women. medRxiv Prepr Serv Heal Sci. 2020. doi:10.1101/2020.07.08.20149179

35. Abrams EM, Szefler SJ. COVID-19 and the impact of social determinants of health. Lancet Respir Med. 2020;8(7):659–661. doi:10.1016/S2213-2600(20)30234-4

36. Janevic T, Zeitlin J, Egorova N, Hebert PL, Balbierz A, Howell EA. Neighborhood Racial And Economic Polarization, Hospital Of Delivery, And Severe Maternal Morbidity. https://doi.org/101377/hlthaff201900735. May 2020. doi:10.1377/HLTHAFF.2019.00735

37. Tangel V, White RS, Nachamie AS, Pick JS. Racial and Ethnic Disparities in Maternal Outcomes and the Disadvantage of Peripartum Black Women: A Multistate Analysis, 2007-2014. Am J Perinatol. 2019;36(8):835–848. doi:10.1055/s-0038-1675207

38. Grobman WA, Parker CB, Willinger M, et al. Racial disparities in adverse pregnancy outcomes and psychosocial stress. Obstet Gynecol. 2018;131(2):328–335. doi:10.1097/AOG.0000000000002441

39. Crawford S, Joshi N, Boulet SL, et al. Maternal Racial and Ethnic Disparities in Neonatal Birth Outcomes with and Without Assisted Reproduction. Obstet Gynecol. 2017;129(6):1022–1030. doi:10.1097/AOG.0000000000002031

40. Marin Gabriel MA, Vergeli MR, Caserio Carbonero S, et al. Maternal, Perinatal and Neonatal Outcomes With COVID-19: A Multicenter Study of 242 Pregnancies and Their 248 Infant Newborns During Their First Month of Life. Pediatr Infect Dis J. 2020.

41. Egerup P, Olsen LF, Christiansen A-MH, et al. Severe Acute Respiratory Syndrome Coronavirus 2 (SARS-CoV-2) Antibodies at Delivery in Women, Partners, and Newborns. Obstet Gynecol. 2020.

42. Atladóttir HÓ, Thorsen P, Østergaard L, et al. Maternal infection requiring hospitalization during pregnancy and autism spectrum disorders. J Autism Dev Disord. 2010;40(12):1423–1430. doi:10.1007/s10803-010-1006-y

43. Meyer U, Nyffeler M, Yee BK, Knuesel I, Feldon J. Adult brain and behavioral pathological markers of prenatal immune challenge during early/middle and late fetal development in mice. Brain Behav Immun. 2008;22(4):469–486. doi:10.1016/j.bbi.2007.09.012

44. Meyer U, Nyffeler M, Engler A, et al. The time of prenatal immune challenge determines the specificity of inflammation-mediated brain and behavioral pathology. J Neurosci. 2006;26(18):4752–4762. doi:10.1523/JNEUROSCI.0099-06.2006

